# Validity of self-testing at home with rapid SARS-CoV-2 antibody detection by lateral flow immunoassay

**DOI:** 10.1101/2022.06.08.22276154

**Authors:** Christina J Atchison, Maya Moshe, Jonathan C Brown, Matthew Whitaker, Nathan C K Wong, Anil A Bharath, Rachel A McKendry, Ara Darzi, Deborah Ashby, Christl A. Donnelly, Steven Riley, Paul Elliott, Wendy S Barclay, Graham S Cooke, Helen Ward

## Abstract

**Background:** Severe acute respiratory syndrome coronavirus 2 (SARS-CoV-2) antibody lateral flow immunoassays (LFIA) can be carried out in the home and have been used as an affordable and practical approach to large-scale antibody prevalence studies. However, assay performance differs from that of high-throughput laboratory-based assays which can be highly sensitive. We explore LFIA performance under field conditions compared to laboratory-based ELISA and assess the potential of LFIAs to identify people who lack functional antibodies following infection or vaccination.

**Methods:** Field evaluation of a self-administered LFIA test (Fortress, NI) among 3758 participants from the REal-time Assessment of Community Transmission-2 (REACT-2) study in England selected based on vaccination history and previous LFIA result to ensure a range of antibody titres. In July 2021, participants performed, at home, a self-administered LFIA on finger-prick blood, reported and submitted a photograph of the result, and provided a self-collected capillary blood sample (Tasso-SST) for serological assessment of IgG antibodies to the spike protein using the Roche Elecsys® Anti-SARS-CoV-2 assay. We compared the self-administered and reported LFIA result to the quantitative Roche assay and checked the reading of the LFIA result with an automated image analysis (ALFA). In a subsample of 250 participants, we compared the results to live virus neutralisation.

**Results:** Almost all participants (3593/3758, 95.6%) had been vaccinated or reported prior infection, with most having received one (862, 22.9%) or two (2430, 64.7%) COVID-19 vaccine doses. Overall, 2777/3758 (73.9%) were positive on self-reported LFIA, 2811/3457 (81.3%) positive by LFIA when ALFA-reported, and 3622/3758 (96.4%) positive on Roche anti-S (using the manufacturer reference standard threshold for positivity of 0.8 U ml^-1^). Live virus neutralisation was detected in 169 of 250 randomly selected samples (67.6%); 133/169 were positive with self-reported LFIA (sensitivity 78.7%; 95% CI 71.8, 84.6), 142/155 (91.6%; 86.1, 95.5) with ALFA, and 169 (100%; 97.8, 100.0) with Roche anti-S. There were 81 samples with no detectable virus neutralisation; 47/81 were negative with self-reported LFIA (specificity 58.0%; 95% CI 46.5, 68.9), 34/75 (45.3%; 33.8, 57.3) with ALFA, and 0/81 (0%; 0.0, 4.5) with Roche anti-S. All 250 samples remained positive with Roche anti-S when the threshold was increased to 1000U ml^-1^.

**Conclusions:** Self-administered LFIA can provide insights into population patterns of infection and vaccine response, and sensitivity can be improved with automated reading of the result. The LFIA is less sensitive than a quantitative antibody test, but the positivity in LFIA correlates better than the quantitative ELISA with virus neutralisation.

## Introduction

In April 2020 the UK Government initiated a programme of at-home SARS-CoV-2 antibody testing using self-administered finger prick lateral flow immunoassays (LFIAs) to provide community prevalence estimates of antibodies to SARS-CoV-2 in England (1). As part of this programme, the REal-time Assessment of Community Transmission-2 (REACT-2) study used this approach to estimate the number and distribution of infections during the first wave of the COVID-19 pandemic (2), identify the decrease in antibody positivity over time (3) and measure the impact of vaccine roll-out on population antibody prevalence (4). As such, REACT-2 has been able to show that LFIAs used at home for self-testing are an affordable and practical approach to large-scale antibody testing for SARS-CoV-2 antibody prevalence, providing rapid results without the support of central laboratories (3). As COVID-19 vaccination programmes are rolled out worldwide, population antibody testing could have an important additional role in monitoring immune responses to vaccinations, informing policy regarding booster doses, and assessing levels of potentially protective immunity in the population (5).

The REACT-2 programme conducted extensive clinical and laboratory evaluation of SARS-CoV-2 antibody LFIA performance by sampling from healthcare workers (6-8), non-healthcare key workers (9) and patients from a renal transplant cohort (8), summarised in Supplementary Table S1. High acceptability and usability for self-testing with LFIAs was demonstrated among non-healthcare key workers and a random sample of adults in the population (9, 10). The LFIA selected for the REACT-2 large-scale antibody prevalence study (Fortress, Northern Ireland) detects antibody against the spike (“S”) protein of the virus (contained in, or coded by, all UK licensed vaccines). It was initially evaluated in a healthcare worker cohort known to have been infected with SARS-CoV-2, with a sensitivity 84.0% (95% confidence interval [CI] 70.5, 93.5) and specificity 98.6% (95% CI 97.1, 99.4) (6) and later in a vaccinated population with a sensitivity of 92.3% (95% CI 81.5, 97.9) (Supplementary Table S1).

Prevalence studies based on self-administered LFIA have generally produced a lower estimate of population SARS-CoV-2 antibody positivity than those using quantitative laboratory assays, despite adjustment for test performance (11). As a threshold test, it is likely that the LFIA is predominantly missing people with low antibody titres. The clinical significance of detectable but low antibody titres (post-infection or post-vaccine) picked up by quantitative laboratory assays is unclear but is likely to correlate with lower protection from infection (12). Calibrated to the appropriate positivity threshold for protection, the LFIA could be a feasible tool for monitoring the distribution of protective serological antibody responses in the population and could be useful as a screening tool for identifying individuals with below threshold antibody levels who may benefit from further vaccination or other prevention measures. To further explore this hypothesis and investigate the utility of the Fortress LFIA under field conditions, we compare results of self-reported qualitative LFIA results against a quantitative laboratory-based ELISA performed on simultaneously self-collected capillary blood. Given the strong evidence of a protective role for neutralising serum antibodies (13, 14), and evidence for correlation between SARS-CoV-2 IgG antibody values and neutralisation titres (15), we also explore the relationship between LFIA results and antibody titres with viral neutralisation.

## Methods

### Study design and sampling

This was a field evaluation study conducted between 1^st^ July 2021 and 10^th^ August 2021.

This study recruited participants from round 6 of the national REACT-2 study of SARS-CoV-2 antibody prevalence in England who had given permission to be contacted for further research. Methods for the national REACT-2 study, including round 6, are published elsewhere (1, 16). Briefly, REACT-2 is a series of cross-sectional population surveys of the prevalence of SARS-CoV-2 antibodies in the community in England, UK. At each round, we contacted a random sample of the population by sending a letter to named individuals aged 18 or over from the National Health Service (NHS) patient list (covering almost the whole population) and respondents were sent an LFIA self-testing kit to perform at home.

For this follow-up study, purposeful random sampling was carried out by re-contacting 7000 participants who had participated in round 6 of REACT-2 in May 2021, aiming to achieve a sample size of 4000. We invited equal numbers in each of the following categories based on results from round 6 – unvaccinated and LFIA negative, double vaccinated (>20 days previously) and LFIA negative, unvaccinated and LFIA positive, and double vaccinated and LFIA positive. This sampling frame was chosen to recruit sufficient people with positive and negative self-test results post-infection and post-vaccination, recognising that many people would have received further vaccination in the interim.

People were invited by post to register until approximately 4000 had signed up. Registration was undertaken online or by telephone. Those who registered were sent a further LFIA test kit to carry out at home, and asked to report the result online, upload a photograph of the result, and complete a short online questionnaire. In addition, participants were asked to take a 400 to 500μl capillary blood sample at the same time-point using an at-home self-collection blood device (Tasso-SST (17)) and return the sample for serological assessment of antibodies. Participants were offered an incentive of £20 on completion of all study tasks.

### ALFA (Automated Lateral Flow Analysis): machine learning algorithm for automated analysis of LFIA images

The national REACT-2 study was based on self-reported LFIA results, meaning that prevalence estimates rely on subjective interpretation by participants of a test line by eye, risking false positives and negatives. However, we have shown in previous evaluations that participant reported LFIA interpretation is consistent with clinician interpreted results (9, 10). We went further and developed a computational pipeline (ALFA) which used machine learning algorithms to analyse participant-submitted images of the Fortress LFIA from REACT-2 rounds 1 to 5, and subsequently classify results as invalid, IgG negative and IgG positive. Methods for development of ALFA are published elsewhere (in press). Automated analysis showed substantial agreement with human experts and performed consistently better than study participants, particularly for weak positive IgG results. Our findings supported the use of machine learning-enabled automated reading of at-home antibody lateral flow tests as a tool for improved accuracy (in press).

### Laboratory Methods

Serological assessment was performed in a commercial laboratory on the Roche Elecsys® Anti-SARS-CoV-2 assay which reports a quantitative anti-Spike (anti-S) antibody titre. This assay has been previously validated by Public Health England who reported a specificity of 100% (95% CI 99.1, 100), and an overall sensitivity of 98.5% (95% CI 96.9, 99.4) in samples 21 days post-onset in people with PCR-confirmed infection (18). The threshold value for antibody positivity for the Roche assay is 0.8 U ml^-1^ based on manufacturer instructions (18). The lower limit of quantification is 0.4 U ml^-1^ (19). Measurements below this value were truncated at 0.4 U ml^-1^. The assay was analysed in its original scale (U ml^-1^). WHO international standard units are BAU ml^-1^ for anti-spike IgG to allow comparison across studies and platforms (20). The conversion factor for U ml^-1^ to BAU ml^-1^ for the Roche Elecsys® Anti-SARS-CoV-2 assay:

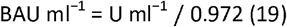

In addition, we selected 250 serum samples at random for assessment on a live virus neutralisation assay. Serum samples were heat-inactivated and a 2-fold dilution series was performed in 96-well plates. Serum dilutions were incubated with 100 TCID_50_ SARS-CoV-2 (WT D614G) for 1 hour at 37°C. Vero E6 cells modified to overexpress ACE2 and TMPRSS2 (VAT cells) were then added to the wells and incubated at 37°C for 72 hours before assessing the cells for the presence or absence of virus-induced cytopathic effect (CPE). The neutralisation titre of a serum sample was defined as the reciprocal of the highest serum dilution at which CPE was not observed, demonstrating antibody-mediated protection from virus – e.g. protection of cells at a 1:20 dilution of serum gives a neutralisation titre value of 20. Serum samples were titrated 2-fold in duplicate with a starting dilution of 1:10 meaning that if 1 of the 2 replicate wells were protected at this first dilution, the titre was expressed as 7.1, halfway to the 1:10 dilution on a log2 scale. Serum samples for which CPE was observed in all wells were therefore defined as having neutralisation titre of <7.1. Serum samples were run alongside a WHO antibody reference standard (antibody 20/150, NIBSC) which scored a neutralisation titre of 320 on our assay. This standard has a defined potency of 832 BAU ml^-1^ anti-Spike IgG which allowed for indirect conversion of the neutralisation titres of serum samples to BAU ml^-1^ (21) (Supplementary Figure S1). Using a calculated conversion factor of 2.6 BAU per neutralisation titre unit, the lower limit of detection of 7.1 equates to 18.5 BAU ml^-1^.

### Data analysis

We report on the positivity based on three results for each participant: self-administered and reported LFIA (hereafter self-LFIA), self-administered and machine-read LFIA (hereafter ALFA) and Roche Elecsys® platform (hereafter ELISA) using the manufacturer recommended threshold ≥0.8 U ml^-1^. As the manufacturer’s threshold for antibody positivity for the Roche assay is likely too low to correlate with moderate-to-high levels of protection from infection based on recent studies in the UK population (12, 15), we also report positivity at different thresholds of ≥100 U ml^-1^, ≥350 U ml^-1^ and ≥1000 U ml^-1^ – equivalent to ≥103 BAU ml^-1^, ≥360 BAU ml^-1^ and ≥1029 BAU ml^-1^, respectively. In addition, we report the distribution of quantitative ELISA results for self-LFIA and ALFA positive and negative results.

Results are presented with the corresponding binomial exact 95% confidence interval (95% CI). Quantitative variables were described as medians and interquartile ranges (IQR). We also determined the geometric mean titre (GMT) for anti-S antibody. Categorical variables were presented as frequencies and percentage.

We assessed the association between self-LFIA, ALFA, ELISA and live virus neutralisation titres, with the threshold of neutralisation detection defined as a titre of ≥7.1 (equivalent to 18.5 BAU ml^-1^, see methods). We then used this as a standard to determine sensitivity and specificity of self-LFIA, ALFA and ELISA at different thresholds as a measure of neutralisation. The non-parametric Mann-Whitney test was performed to compare neutralisation titres according to whether positive or negative by self-LFIA, and to compare IgG antibody titres according to whether positive or negative by self-LFIA.

We used multiple linear regression to quantify associations between demographic characteristics, history of COVID-19, vaccination status and time since double vaccinated (two doses) and log_10_-transformed antibody titres. For the interpretation of the regression coefficients in the models, where only the dependent variable is log-transformed, the coefficient is exponentiated, one subtracted from this number, and multiplied by 100. This gives the estimated percent increase (or decrease) in antibody titres for every one-unit increase in the independent variable. Example: if the coefficient is 0.198, (exp(0.198) – 1) * 100 = 21.9. Thus, for every one-unit increase in the independent variable, antibody titre is estimated to increase by about 22%.

Data were analysed using the statistical packages STATA version 15.0 and GraphPad Prism 9.0.0. The threshold for statistical significance was p <0.05 (2-sided).

### Patient and public involvement

Public involvement and participant feedback were central to the design of the REACT-2 programme. There has been extensive involvement from patient panels and rigorous evaluation of the usability of LFIAs included in the REACT-2 studies.

#### Ethics

This work was undertaken as part of the REACT-2 study, with ethical approval from South Central–Berkshire B Research Ethics Committee (REC ref: 20/SC/0206; IRAS 283805).

## Results

Overall, 71.0% (4972/7000) of invited individuals agreed to take part in the study, of whom, 1214 (24.4%) were excluded from the analysis due to either a missing or invalid self-LFIA result (n=327) or a missing or void ELISA result (n=887). The reasons for the large number of missing or void ELISA results include insufficient and incorrectly labelled samples and laboratory error, but the distribution of these was not provided by the commercial laboratory performing the tests. A total of 3758 participants had paired self-LFIA and ELISA results, 96.6% (3457/3578) of whom also uploaded a photograph of their self-LFIA test which enabled analysis using the ALFA algorithm. Participant characteristics are shown in Table 1. The majority of the participants had received one (862, 22.9%) or two (2430, 64.7%) COVID-19 vaccine doses, and 27.4% reported suspected or confirmed past COVID-19 (Table 1), meaning that almost all participants (3593/3758, 95.6%) reported either vaccine or prior infection.

**Table 1:**
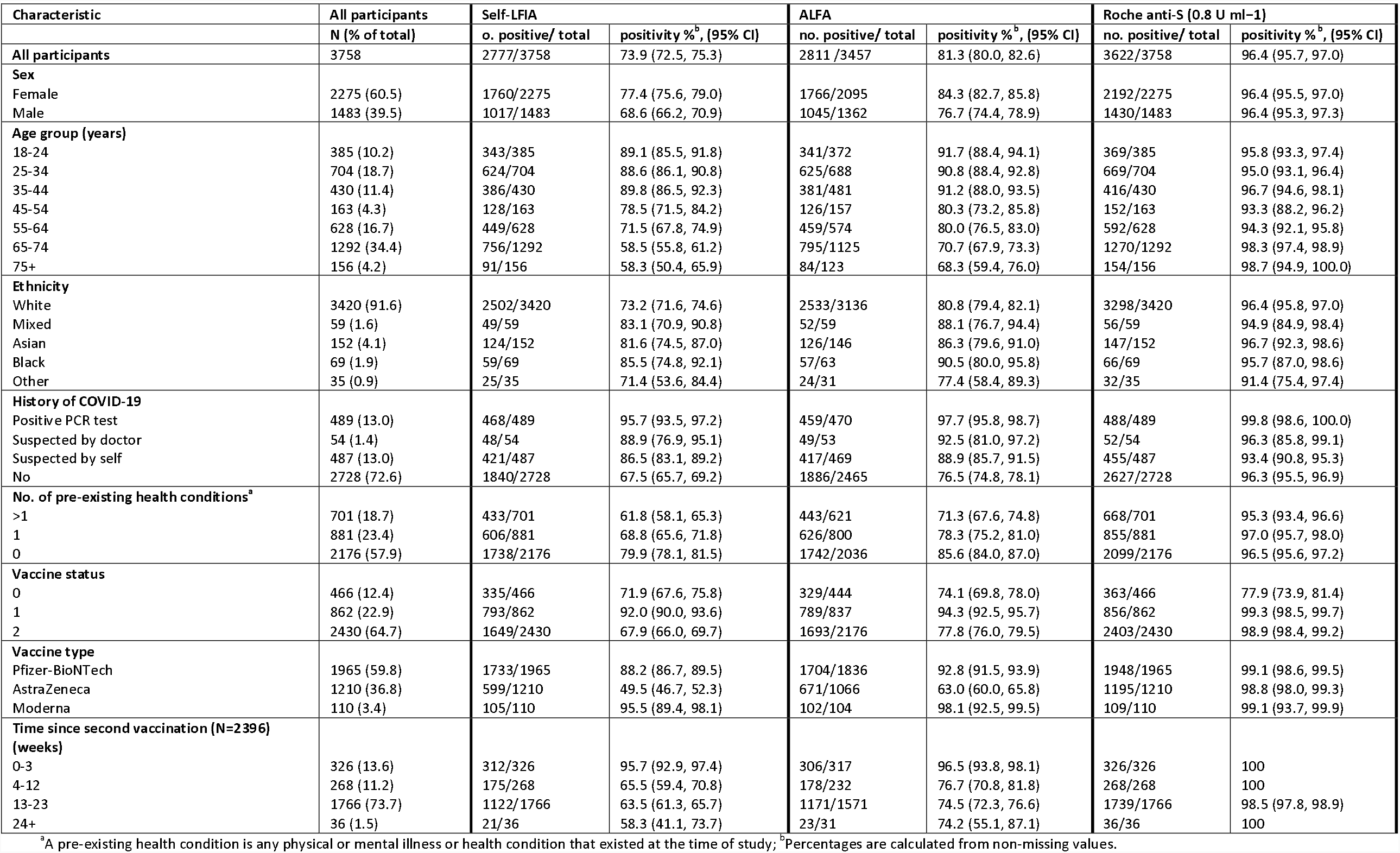
Demographic and clinical characteristics of the study participants by antibody status for self-LFIA, ALFA and Roche anti-S at 0.8U ml^-1^.

### IgG anti-S positivity and antibody titres

Self-LFIA positivity was 73.9% (2777/3758, 95% CI 72.5, 75.3) (Table 1); ALFA positivity was 81.3% (2811/3457, 95% CI 80.0, 82.6), and ELISA positivity was 96.4% (3622/3758, 95% CI 95.7, 97.0) using the manufacturer’s threshold of ≥0.8 U ml^-1^. ELISA positivity decreased to 83.1% (95% CI 81.9, 84.3), 62.7% (95% CI 61.1, 64.2) and 47.0% (95% CI 45.4, 48.6) by increasing the ELISA threshold to ≥100 U ml^-1^, ≥350 U ml^-1^ and ≥1000 U ml^-1^, respectively.

Figure 1 shows the distribution of ELISA titres for samples that were positive and negative on self-reported LFIA. The self-LFIA positive samples had a median anti-S titre of 1702.0 U ml^-1^ (IQR 357.9 to 7416.0) and a range of 0.40 U ml^-1^ to 25000.0 U ml^-1^. The self-LFIA negative samples had a median anti-S titre of 142.6 U ml^-1^ (IQR 46.6 to 384.0). There were 859 discrepant results with a negative self-LFIA and a positive ELISA; for these samples the median anti-S titre was 197.6 U ml^-1^ (IQR 78.9 to 443.7) indicating that these were weaker positives on average. Of the self-LFIA positive samples with a negative Roche assay (n=14), the median anti-S titre was 0.4 U ml^-1^; anti-S titre ranged from 0.4 U ml^-1^ to 0.75 U ml^-1^ indicating false positives (Table 2).

**Table 2:** Comparison of results from paired self-LFIA and ALFA, and Roche anti-S (using the manufacturer’s threshold of ≥0.8 U ml^-1^), N=3758.

| Self-LFIA | Roche positive<br>N<br>(median (IQR) titre) | Roche negative<br>N<br>(median (IQR) titre) | Total<br>N<br>(median (IQR) titre) |
| --- | --- | --- | --- |
| **Positive** | 2763<br>(1715.0; 368.9-7489.0) | 14<br>(0.4; 0.4-0.4) | 2777<br>(1702.0; 357.9-7416.0) |
| **Negative** | 859<br>(197.6; 78.9-443.7) | 122<br>(0.4; 0.4-0.4) | 981<br>(142.6; 46.6-384.0) |
| **Total** | 3622<br>(925.4; 207.5-4655.0) | 136<br>(0.4; 0.4-0.4) | 3758<br>(824.1; 168.5-4286.0) |
| **ALFA** |  |  |  |
| **Positive** | 2798<br>(1566.5; 313.0-7119.0) | 13<br>(0.4; 0.4-0.4) | 2811<br>(1541.0; 306.2-7079.0) |
| **Negative** | 531<br>(131.6; 63.3-267.3) | 115<br>(0.4; 0.4-0.4) | 646<br>(102.7; 24.7-235.7) |
| **Total** | 3329<br>(947.4; 201.4-4990.0) | 128<br>(0.4; 0.4-0.4) | 3457<br>(831.5; 165.1-4668.0) |

**Figure 1:**
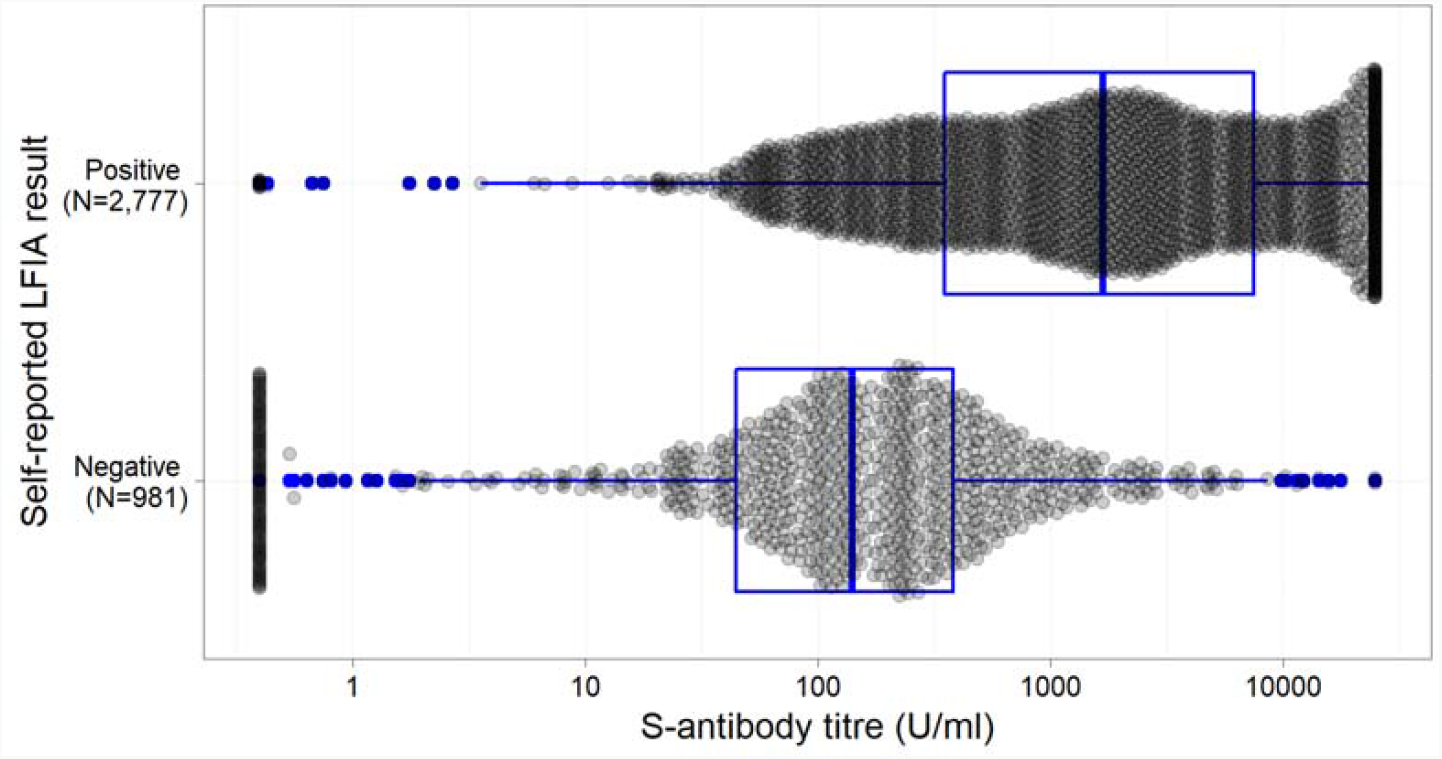
Box plot (median and quartiles) illustrating the distribution of quantitative Roche anti-S titres by self-LFIA result (N=3758)

Table 2 also shows the comparison using the machine-read (ALFA) LFIA results; for samples with a negative ALFA and positive ELISA, the median anti-S titre was lower than self-LFIA at 131.67 (IQR 63.3-267.3) suggesting that ALFA was better at detecting weaker positives.

Supplementary Table S2 shows the same results calibrated with anti-S thresholds of ≥100 U ml^-1^, ≥350 U ml^-1^ and ≥1000 U ml^-1^.

### Live Virus Neutralisation

Neutralisation assays were performed on 250 randomly selected serum samples, including 167 self-reported positive and 83 self-reported negative LFIA participants.

Live virus neutralisation was detected in 169 of 250 samples. The self-LFIA had an estimated sensitivity of 78.7% (133/169; 95% CI 71.8, 84.6) and specificity of 58.0% (47/81; 95% CI 46.5, 68.9) using detectable neutralisation (equivalent to at least 18.5 BAU ml^-1^) as the comparator (Table 3). The ALFA-LFIA had an estimated sensitivity of 92.3% (142/155; 95% CI 86.9, 95.9) and specificity of 45.3% (34/75; 95% CI 33.8, 57.3) (Table 3). The ELISA had a sensitivity of 100% (95% CI 97.8, 100.0) and specificity of 0% (95% CI 0.0, 4.5) as all neutralisation titres <7.1 threshold were positive on the Roche assay (Table 3). All 250 samples remained positive by ELISA when the anti-S titre threshold was increased to 1000 U ml^-1^.

**Table 3:** Comparison of results from self-LFIA and ALFA, and SARS-CoV-2 neutralisation titre (NT) (neutralisation titres of 7.1 have been assigned an arbitrary threshold of 0.1), N=250 (Self-LFIA) and N=230 (ALFA).

| Self- LFIA | NT positive<br>(median (IQR) titre) | NT negative<br>(median (IQR) titre) | Total<br>(median (IQR) titre) | Performance (95% CI) |
| --- | --- | --- | --- | --- |
| **Positive** | 133<br>(20.0; 10.0-113.1) | 34<br>(0.1; 0.1-0.1) | 167<br>(14.1; 7.1-80.0) | Sensitivity: 78.7 (71.8-84.6) |
| **Negative** | 36<br>(10.0; 7.1-14.1) | 47<br>(0.1; 0.1-0.1) | 83<br>(0.1; 0.1-10.0) | Specificity: 58.0 (46.5-68.9) |
| **Total** | 169<br>(20.0; 10.0-80.0) | 81<br>(0.1; 0.1-0.1) | 250<br>(10.0; 0.1-28.3) |  |
| **ALFA** |  |  |  |  |
| **Positive** | 142<br>(20.0; 10.0-104.8) | 41<br>(0.1; 0.1-0.1) | 183<br>(14.1; 7.1-56.6) | Sensitivity: 91.6 (86.1-95.5) |
| **Negative** | 13<br>(10.0; 7.1-14.1) | 34<br>(0.1; 0.1-0.1) | 47<br>(0.1; 0.1-7.1) | Specificity: 45.3 (33.8-57.3) |
| **Total** | 155<br>(20.0; 10.0-80.0) | 75<br>(0.1; 0.1-0.1) | 230<br>(10.0; 0.1-28.3) |  |
| **Roche**<br>**(≥0.8 U ml<sup>-1</sup>)** |  |  |  |  |
| **Positive** | 169<br>(20; 10-80) | 81<br>(0.1; 0.1-0.1) | 250<br>(10; 0.1-28.3) | Sensitivity: 100% (97.8-100) |
| **Negative** | 0<br>- | 0<br>- | 0<br>- | Specificity: 0% (0-4.5) |
| **Total** | 169<br>(20; 10-80) | 81<br>(0.1; 0.1-0.1) | 250<br>(10; 0.1-28.3) |  |

Figure 2 shows the distribution of live virus neutralisation titres against anti-S titres, with points labelled for LFIA positive and negative. Neutralisation titres were higher in participants with positive compared to negative LFIA results (p<0.0001). A similar association was observed for anti-S titres and LFIA result (p<0.0001).

**Figure 2:**
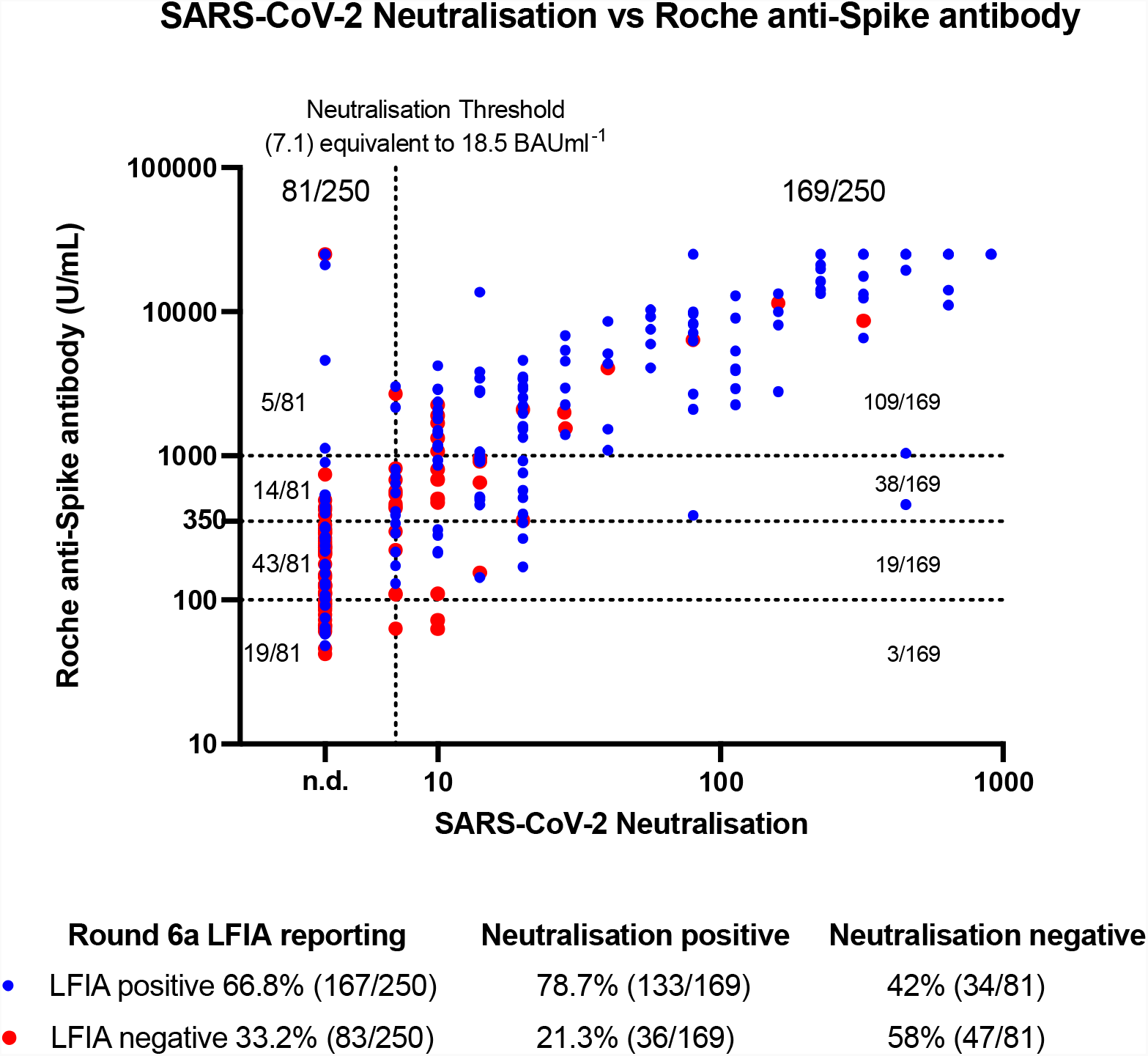
Relationship between SARS-CoV-2 live virus neutralisation titre and the Roche anti-Spike ELISA-based assay by self-LFIA. Positive self-LFIA results are represented in blue and negative LFIA results are represented in red. The threshold of SARS-CoV-2 neutralisation detection is defined as ≥7.1, equivalent to 18.5 BAU ml^-1^, as denoted by the vertical black dotted line and samples below this are marked as not detected (n.d.) Both axes use a Log 10 scale. Roche anti-Spike antibody thresholds of ≥100 U ml^-1^, ≥350 U ml^-1^ and ≥1000 U ml^-1^ are denoted by horizontal dotted lines. Statistical significance is reported by performing a non-parametric Mann-Whitney test for neutralisation titres by self-LFIA positive and negative results (p= 0.0001), and for Roche anti-Spike antibody titres by self-LFIA positive and negative results (p=0.0001).

The conversion of neutralisation titres to BAU ml^-1^ following titration of a WHO antibody reference standard showed that 34.9% (59/169) of the neutralisation positive samples had a titre of ≥100 BAU ml^-1^ (Supplementary Figure 1).

### Factors associated with antibody titres

We found associations between demographic characteristics, history of COVID-19 and vaccination status with levels of SARS-CoV-2 antibody, in keeping with previous studies (12, 22-25). The full results are shown in Supplementary Table S3.

## Discussion

The self-administered Fortress LFIA offers a validated qualitative tool that provides a means for obtaining community-wide SARS-CoV-2 antibody positivity prevalence estimates rapidly and at scale, at reasonable cost by adjusting the results for known test performance. The threshold for positivity of the LFIA is higher than that of laboratory-based ELISA anti-S assays, producing lower estimates of population antibody prevalence.

Although the Fortress LFIA has a threshold that means it does not detect a proportion of positive anti-spike IgG registered on the Roche assay, that threshold is close to the level at which neutralising antibody can be reliably measured. Indeed, we demonstrated that the estimated specificity of the self-administered self-reported Fortress LFIA against positive neutralisation titres was substantially higher than that of the Roche anti-S assay with manufacturer’s threshold of 0.8 U ml^-1^ (58.0% vs. 0%). There is evidence that the presence of neutralising antibodies in sera is highly predictive of protection from symptomatic disease following SARS-CoV-2 infection and that declining levels of neutralising antibody titres correlate with increased risk of symptomatic infection and severe disease (14).

The LFIA is predominantly missing people with low antibody titres, but the implications of a higher threshold for IgG detection on LFIA testing are not yet well understood, and may represent an important marker of protection against SARS-CoV-2 infection and/or severe disease. We question the clinical and epidemiological significance of detectable but low antibody titres picked up by the low thresholds for positivity used for quantitative laboratory assays and suggest that these cut-offs may need to be recalibrated (upwards) to be a useful marker of protection from infection and/or severe disease. Wei at al. recently explored the association between anti-spike IgG levels and protection from SARS-CoV-2 infection with majority Delta (B.1.617.2) variant in a large representative sample of households with longitudinal follow-up (12). They showed that protection against infection rose sharply as antibody levels increased in unvaccinated participants with prior infection, with 67% protection at 33 BAU ml^-1^ using the OmniPATH 384 Combi SARS-CoV-2 IgG ELISA (Thermo Fisher Scientific) assay. Higher antibody levels were required to reach the same level of protection after vaccination, with 67% protection at 107 BAU ml^-1^ or 94 BAU ml^-1^ with ChAdOx1 (Oxford-AstraZeneca) or BNT162b2 (Pfizer), respectively (12). The threshold for determining IgG positivity for the assay used was ≥23 BAU ml^-1^ (12). Similarly, Fent et al. showed a vaccine efficacy of 80% against symptomatic infection with majority Alpha (B.1.1.7) variant was achieved with 264 BAU ml^-1^ (15).

These findings suggest that antibody positivity on the LFIA could be useful to measure any waning of vaccine induced immunity in the population, indeed more useful than quantitative assays with low thresholds for positivity which would result in false reassurance by having higher false positive results. Rapid antibody testing may prove useful in ongoing population surveillance to inform policy for subsequent vaccination programmes, including the targeting of booster vaccines, and has potential for initial screening of patients in the community to receive anti-viral therapy as laboratory-based methods may cause a delay in initiating treatment.

We found that using an in-house machine learning algorithm to read the LFIA results based on images submitted by participants increases detection of antibody, particularly in those with lower anti-S titres, suggesting that participants with weak positive findings may have inaccurately recorded a positive test as negative. This finding was not unexpected as we have previously demonstrated that automated analysis showed substantial agreement with human experts and performed consistently better than study participants, particularly for weak positive IgG results (in press). Thus, this study lends further support for the use of machine learning-enabled automated reading to achieve a gain in accuracy in large-scale community surveys using at-home self-administered LFIAs.

### Strengths and Limitations

Unlike previous evaluations of the Fortress LFIA, this study replicates the ‘real-world’ application of LFIAs in large-scale antibody prevalence studies of the general population where users are self-administering the test in their own homes following detailed instructions. Therefore, the study authentically explores the accuracy of the Fortress LFIA under the field conditions in which it is most likely to be deployed for surveillance.

Our purposeful sampling strategy of selecting approximately equal numbers of unvaccinated and LFIA negative, double vaccinated and LFIA negative, unvaccinated and LFIA positive, and double vaccinated and LFIA positive may have introduced biases. By purposive selection of vaccinated LFIA negative individuals there is the possibility that we enriched our sample for low level antibody titres that might be less common at population level, thus overall figures on sensitivity cannot be extrapolated to real world use in a random population sample.

We compared results of the self-administered Fortress LFIA with a quantitative ELISA test (Roche) using an established cut-point to denote positivity. To the extent that the “gold standard” ELISA test itself does not have perfect sensitivity and specificity (18), its use will have introduced some error into those comparisons.

We used data from 1^st^ July 2021 to 10^th^ August 2021-that is, while the Delta (B.1.617.2) variant accounted for nearly all cases (26). Our neutralisation assays used a first wave isolate as target, with antigenicity the same as the Wuhan strain. In settings in which Delta is not the dominant variant causing disease, or where neutralisation assays use different strains of the virus, the relationships between IgG antibody positivity by LFIA or quantitative anti-S assays and neutralisation titres shown here may not apply. Indeed, Wall et al. demonstrated neutralizing antibody titres were 5.8-fold lower against Delta relative to the Wuhan variant after two doses of BNT162b2 (27). It has also been reported that neutralizing antibody titres for the ChAdOx1 and BNT162b2 vaccines are reduced with the Beta (B.1.351) variant by 9-fold and 7.6-fold, respectively compared with the early Wuhan-related Victoria variant (28). Neutralizing antibody titres against Omicron (B.1.1.529) have been shown to be eight-fold lower than with Delta after two BNT162b2 vaccinations (29). As such, emerging viral variants might need higher antibody levels for the same level of neutralising activity (14). In the case where relationships between antibody levels and levels of protection do not change with other variants and assuming that neutralisation is a major mechanism of protection (or that the mechanism of protection remains correlated with neutralisation over time), future LFIAs could be calibrated to the appropriate antibody positivity threshold for protection.

## Conclusion

In summary, at-home self-testing and reporting with LFIAs provide a rapid and cost-effective means to assess population antibody prevalence of SARS-CoV-2. Gains in accuracy, and thus improvements in prevalence estimates, can be achieved by using machine learning algorithms to read the LFIA results based on images provided by participants. In the future, calibrating the threshold for antibody positivity of LFIAs to binding or neutralising antibody levels correlated with protection from infection and/or severe disease, could provide a valuable role for home-testing by LFIA to inform vaccination and treatment strategies going forward. As a first step it would be important to understand the extent to which a positive LFIA result is predictive of protection against infection, illness and hospitalisation.

## Data Availability

All data produced in the present study are available upon reasonable request to the authors

## Author contributions

HW, CJA and GSC conceptualized and designed the study and drafted the manuscript. CJA, HW, MW, MM, JCB, NCKW, AAB and WSB undertook data collection and data analysis. DA provided statistical advice. HW, GSC, WSB, PE, CAD, SR and AD provided study oversight. AD and PE obtained funding. SR, RAM, AAB, DA, WSB, CAD, AD and PE critically reviewed the manuscript. All authors read and approved the final version of the manuscript. HW is the guarantor for this paper. The corresponding author attests that all listed authors meet authorship criteria and that no others meeting the criteria have been omitted, had full access to all the data in the study, and had final responsibility for the decision to submit for publication.

## Acknowledgments

The authors thank key collaborators on this work— Imperial College London: Eric Johnson and Graham Blakoe. Ipsos: Stephen Finlay, John Kennedy, Duncan Peskett, Sam Clemens and Kelly Beaver; and the REACT Public Advisory Panel.

## References

1. Riley S, Atchison C, Ashby D, Donnelly CA, Barclay W, Cooke GS, et al. REal-time Assessment of Community Transmission (REACT) of SARS-CoV-2 virus: Study protocol. Wellcome Open Res. 2020;5:200.

2. Ward H, Atchison C, Whitaker M, Ainslie KEC, Elliott J, Okell L, et al. SARS-CoV-2 antibody prevalence in England following the first peak of the pandemic. Nature Communications. 2021;12(1):905.

3. Ward H, Cooke GS, Atchison C, Whitaker M, Elliott J, Moshe M, et al. Prevalence of antibody positivity to SARS-CoV-2 following the first peak of infection in England: Serial cross-sectional studies of 365,000 adults. Lancet Reg Health Eur. 2021;4:100098.

4. Ward H, Cooke G, Whitaker M, Redd R, Eales O, Brown JC, et al. REACT-2 Round 5: increasing prevalence of SARS-CoV-2 antibodies demonstrate impact of the second wave and of vaccine roll-out in England. medRxiv. 2021:2021.02.26.21252512.

5. Maple PAC. Population (Antibody) Testing for COVID-19—Technical Challenges, Application and Relevance, an English Perspective. Vaccines. 2021;9(6).

6. Flower B, Brown JC, Simmons B, Moshe M, Frise R, Penn R, et al. Clinical and laboratory evaluation of SARS-CoV-2 lateral flow assays for use in a national COVID-19 seroprevalence survey. Thorax. 2020:thoraxjnl-2020-215732.

7. Moshe M, Daunt A, Flower B, Simmons B, Brown JC, Frise R, et al. SARS-CoV-2 lateral flow assays for possible use in national covid-19 seroprevalence surveys (React 2): diagnostic accuracy study. BMJ. 2021;372:423.

8. Cann A, Clarke C, Brown J, Thomson T, Prendecki M, Moshe M, et al. Severe acute respiratory syndrome coronavirus 2 (SARS-CoV-2) antibody lateral flow assay for antibody prevalence studies following vaccination: a diagnostic accuracy study [version 1; peer review: awaiting peer review]. Wellcome Open Res. 2021;6(358).

9. Davies B, Araghi M, Moshe M, Gao H, Bennet K, Jenkins J, et al. Acceptability, Usability, and Performance of Lateral Flow Immunoassay Tests for Severe Acute Respiratory Syndrome Coronavirus 2 Antibodies: REACT-2 Study of Self-Testing in Nonhealthcare Key Workers. Open Forum Infectious Diseases. 2021;8(11):ofab496.

10. Atchison C, Pristerà P, Cooper E, Papageorgiou V, Redd R, Piggin M, et al. Usability and Acceptability of Home-based Self-testing for Severe Acute Respiratory Syndrome Coronavirus 2 (SARS-CoV-2) Antibodies for Population Surveillance. Clinical Infectious Diseases. 2021;72(9):e384–e93.

11. UK_Government. Coronavirus (COVID-19) latest insights: Antibodies 2022 [Available from: https://www.ons.gov.uk/peoplepopulationandcommunity/healthandsocialcare/conditionsanddiseases/articles/coronaviruscovid19latestinsights/antibodies.

12. Wei J, Pouwels KB, Stoesser N, Matthews PC, Diamond I, Studley R, et al. Antibody responses and correlates of protection in the general population after two doses of the ChAdOx1 or BNT162b2 vaccines. Nature Medicine. 2022.

13. McMahan K, Yu J, Mercado NB, Loos C, Tostanoski LH, Chandrashekar A, et al. Correlates of protection against SARS-CoV-2 in rhesus macaques. Nature. 2021;590(7847):630–4.

14. Khoury DS, Cromer D, Reynaldi A, Schlub TE, Wheatley AK, Juno JA, et al. Neutralizing antibody levels are highly predictive of immune protection from symptomatic SARS-CoV-2 infection. Nature Medicine. 2021;27(7):1205–11.

15. Feng S, Phillips DJ, White T, Sayal H, Aley PK, Bibi S, et al. Correlates of protection against symptomatic and asymptomatic SARS-CoV-2 infection. Nature Medicine. 2021;27(11):2032–40.

16. Ward H, Whitaker M, Tang SN, Atchison C, Darzi A, Donnelly CA, et al. Vaccine uptake and SARS-CoV-2 antibody prevalence among 207,337 adults during May 2021 in England: REACT-2 study. medRxiv. 2021:2021.07.14.21260497.

17. Hendelman T, Chaudhary A, LeClair A, van Leuven K, J C, SL F. Self-collection of capillary blood using Tasso-SST devices for Anti-SARS-CoV-2 IgG antibody testing. PLoS One. 2021;16(9):e0255841.

18. Public_Health_England. Evaluation of Roche Elecsys AntiSARS-CoV-2 S serology assay for the detection of anti-SARS-CoV-2 S antibodies 2021 [Available from: https://assets.publishing.service.gov.uk/government/uploads/system/uploads/attachment_data/file/989460/Evaluation_of_Roche_Elecsys_anti_SARS_CoV_2_S_assay_PHE.pdf.

19. Lukaszuk K, Kiewisz J, Rozanska K, Podolak A, Jakiel G, Woclawek-Potocka I, et al. Is WHO International Standard for Anti-SARS-CoV-2 Immunoglobulin Clinically Useful? medRxiv. 2021:2021.04.29.21256246.

20. Infantino M, Pieri M, Nuccetelli M, Grossi V, Lari B, Tomassetti F, et al. The WHO International Standard for COVID-19 serological tests: towards harmonization of anti-spike assays. International Immunopharmacology. 2021;100:108095.

21. Mattiuzzo G, Bentley EM, Hassall M, Routley S, Richardson S, Bernasconi V, et al. WHO/BS.2020.2403 Establishment of the WHO International Standard and Reference Panel for anti-SARS-CoV-2 antibody2020; (20 April 2022). Available from: https://www.nibsc.org/documents/ifu/20-268.pdf.

22. Steensels D, Pierlet N, Penders J, Mesotten D, Heylen L. Comparison of SARS-CoV-2 Antibody Response Following Vaccination With BNT162b2 and mRNA-1273. JAMA. 2021;326(15):1533–5.

23. Zeng F, Dai C, Cai P, Wang J, Xu L, Li J, et al. A comparison study of SARS-CoV-2 IgG antibody between male and female COVID-19 patients: A possible reason underlying different outcome between sex. J Med Virol. 2020;92(10):2050–4.

24. Wei J, Matthews PC, Stoesser N, Maddox T, Lorenzi L, Studley R, et al. Anti-spike antibody response to natural SARS-CoV-2 infection in the general population. Nature Communications. 2021;12(1):6250.

25. Pellini R, Venuti A, Pimpinelli F, Abril E, Blandino G, Campo F, et al. Initial observations on age, gender, BMI and hypertension in antibody responses to SARS-CoV-2 BNT162b2 vaccine. EClinicalMedicine. 2021;36:100928–.

26. UK_Health_Security_Agency. SARS-CoV-2 variants of concern and variants under investigation in England. 2021 [Available from: https://assets.publishing.service.gov.uk/government/uploads/system/uploads/attachment_data/file/1025827/Technical_Briefing_25.pdf.

27. Wall EC, Wu M, Harvey R, Kelly G, Warchal S, Sawyer C, et al. Neutralising antibody activity against SARS-CoV-2 VOCs B.1.617.2 and B.1.351 by BNT162b2 vaccination. Lancet. 2021;397(10292):2331–3.

28. Zhou D, Dejnirattisai W, Supasa P, Liu C, Mentzer AJ, Ginn HM, et al. Evidence of escape of SARS-CoV-2 variant B.1.351 from natural and vaccine-induced sera. Cell. 2021;184(9):2348–61 e6.

29. Nemet I, Kliker L, Lustig Y, Zuckerman N, Erster O, Cohen C, et al. Third BNT162b2 Vaccination Neutralization of SARS-CoV-2 Omicron Infection. New England Journal of Medicine. 2021;386(5):492–4.

