## Supplemental Table 1 for "Validity of self-testing at home with rapid SARS-CoV-2 antibody detection by lateral flow immunoassay"

**Supplementary Table S1: Results of the REACT-2 programme’s diagnostic accuracy evaluations of the Fortress LFIA**

| Study | Participants | Sensitivity (%) (95% CI) | Sample type | Operator | Reference standard(s) | Participants | Sample type | Specificity (%)  (95% CI) |
| --- | --- | --- | --- | --- | --- | --- | --- | --- |
| Flowers et al. (1) | Healthcare workers, N=291 | 88.0 (83.3, 91.2) | Sera | Research technician in laboratory | ^1^S-ELISA and/or hybrid DABA | Police Force, N=500 | Sera from Nov 2019 | 98.6 (97.1, 99.4) |
|  | Healthcare workers, N=45 | 84.0 (70.5, 93.5) | Blood (finger-prick self-test) | Participant in clinic (self-test) | ^1^S-ELISA and/or hybrid DABA |  |  |  |
| Davies et al. (2) | Police Force & Fire Service, N=4705 | 82.1 (77.7, 86.0) | Blood (finger-prick self-test) | Participant in clinic (self-test) | ^2^ELISA (Abbott) |  |  | ^3^97.8 (97.3, 98.2) |
|  | Police Force & Fire Service, N=5206 | 76.4 (71.9, 80.5) | Blood (finger-prick self-test) | Nurse-performed in clinic | ^2^ELISA (Abbott) |  |  | ^3^98.5 (98.1, 98.8) |
| Cann et al. (3) | Vaccinated healthcare workers, N=39 | 92.3 (81.5, 97.9) | Blood (finger-prick self-test) | Participant in clinic (self-test) – result reported by research team | Abbott Architect SARS-CoV-2 IgG Quant II CMIA |  |  | ^3^96.2 (80.4, 99.9) |
|  | Vaccinated renal transplant patients, N=108 | 91.7 (82.7, 96.9) | Blood (finger-prick self-test) | Participant in clinic (self-test) – result reported by research team | Abbott Architect SARS-CoV-2 IgG Quant II CMIA |  |  | ^3^91.7 (77.5, 98.3) |

^1^in-house SARS-CoV-2 spike protein enzyme linked immunoassay (S-ELISA) and a hybrid double antigen binding assay (hybrid DABA), ^2^ELISA (Abbott Laboratories, Lake Bluff, IL, USA) for SARS-CoV-2 immunoglobulin (Ig)G antibody. ^3^LFIA specificity in detecting SARS-CoV-2 IgG antibodies determined using same participants, sample type and ELISA platform as per sensitivity analysis.

**References**

1.	Flower B, Brown JC, Simmons B, Moshe M, Frise R, Penn R, et al. Clinical and laboratory evaluation of SARS-CoV-2 lateral flow assays for use in a national COVID-19 seroprevalence survey. Thorax. 2020:thoraxjnl-2020-215732.

2.	Davies B, Araghi M, Moshe M, Gao H, Bennet K, Jenkins J, et al. Acceptability, Usability, and Performance of Lateral Flow Immunoassay Tests for Severe Acute Respiratory Syndrome Coronavirus 2 Antibodies: REACT-2 Study of Self-Testing in Nonhealthcare Key Workers. Open Forum Infectious Diseases. 2021;8(11):ofab496.

3.	Cann A, Clarke C, Brown J, Thomson T, Prendecki M, Moshe M, et al. Severe acute respiratory syndrome coronavirus 2 (SARS-CoV-2) antibody lateral flow assay for antibody prevalence studies following vaccination: a diagnostic accuracy study [version 1; peer review: awaiting peer review]. Wellcome Open Res. 2021;6(358).
