## Supplemental Table 2 for "Validity of self-testing at home with rapid SARS-CoV-2 antibody detection by lateral flow immunoassay"

**Supplementary Table S2: Comparison of results from paired self-reported LFIA and ALFA, and Roche anti-S (using thresholds of ≥100 U ml^−1^, ≥350 U ml^−1^ and ≥1000 U ml^−1^) N=3758**

| ≥100 U/ml | |  |  |
| --- | --- | --- | --- |
| Self- LFIA | **Roche positive**  **(median (IQR) titre)** | **Roche negative**  **(median (IQR) titre)** | **Total**  **(median (IQR) titre)** |
| Positive | 2529  (2154.0; 574.8-8801.0) | 248  (60.3; 43.9-78.9) | 2777  (1702.0; 357.9-7416.0) |
| Negative | 595  (311.0; 184.6-629.2) | 386  (26.0; 0.4-62.7) | 981  (142.6; 46.6-384.0) |
| Total | 3124  (1429.0; 359.5-6273.0) | 634  (47.3; 5.2-71.0) | 3758  (824.1; 168.5-4286.0) |
| ALFA |  |  |  |
| Positive | 2544  (1975.5; 486.4-8425.5) | 267  (60.4; 44.5-79.0) | 2811  1541.0 (306.2-7079.0) |
| Negative | 328  (233.7; 144.2-396.0) | 318  (22.8; 0.4-59.8) | 646  102.7 (24.7-235.7) |
| Total | 2872  (1488.5; 347.1-6815.5) | 585  (47.8; 3.8-71.0) | 3457  831.5 (165.1-4668.0) |
| ≥350 U/ml | |  |  |
| Self- LFIA | **Roche positive**  **(median (IQR) titre)** | **Roche negative**  **(median (IQR) titre)** | **Total**  **(median (IQR) titre)** |
| Positive | 2089  (3200.0; 1251.0-11768.0) | 688  (137.2; 72.9-224.4) | 2777  (1702.0; 357.9-7416.0) |
| Negative | 267  (704.2; 474.7-1485.0) | 714  (90.8; 22.6-186.0) | 981  (142.6; 46.6-384.0) |
| Total | 2356  (2670.5; 995.2-10181.5) | 1402  (110.7; 52.8-209.0) | 3758  (824.1; 168.5-4286.0) |
| ALFA |  |  |  |
| Positive | 2052  (3140.0; 1162.5-11746.0) | 759  (138.0; 74.8-231.1) | 2811  1541.0 (306.2-7079.0) |
| Negative | 101  (565.2; 438.2-971.2) | 545  (75.0; 8.2-152.1) | 646  102.7 (24.7-235.7) |
| Total | 2153  (2882.0; 1039.0-10941.0) | 1304  (111.0; 53.9-208.4) | 3457  831.5 (165.1-4668.0) |
| ≥1000 U/ml | |  |  |
| Self- LFIA | **Roche positive**  **(median (IQR) titre)** | **Roche negative**  **(median (IQR) titre)** | **Total**  **(median (IQR) titre)** |
| Positive | 1670  (5200.0; 2181.0-15403.0) | 1107  (249.8; 108.9-509.2) | 2777  (1702.0; 357.9-7416.0) |
| Negative | 96  (2215.0; 1388.5-4077.0) | 885  (122.5; 38.2-280.2) | 981  (142.6; 46.6-384.0) |
| Total | 1766  (4790.5; 2114.0-14606.0) | 1992  (190.0; 73.9-401.4) | 3758  (824.1; 168.5-4286.0) |
| ALFA |  |  |  |
| Positive | 1612  (5465.5; 2195.5-15522.5) | 1199  (248.2; 109.1-480.9) | 2811  1541.0 (306.2-7079.0) |
| Negative | 24  (1589.0; 1190.0-2311.5) | 622  (98.9; 19.4-217.4) | 646  102.7 (24.7-235.7) |
| Total | 1636  (5218.0; 2158.5-15387.5) | 1821  (180.9; 73.2-390.8) | 3457  831.5 (165.1-4668.0) |
