## Supplemental Table 3 for "Validity of self-testing at home with rapid SARS-CoV-2 antibody detection by lateral flow immunoassay"

**Supplementary Table S3: Median and geometric mean antibody titres by demographic and clinical characteristics and multivariable linear regression in relation to log_10_-transformed antibody titres**

| **Characteristic** | **Roche anti-S titres**  **(median (IQR))** | **Roche anti-S titres**  **(Geometric Mean (95% CI))** | **Unadjusted regression co-efficient**  **(95% CI)** | **Adjusted regression co-efficient**  **(95% CI)** |
| --- | --- | --- | --- | --- |
| **All participants** | 824.1 (168.5-4286) |  | - | - |
| **Sex**  Female  Male | 942.8 (180.2-4666)  641.2 (154.4-3575) | 768.9 (694.9, 850.6)  643.8 (568.6, 728.9) | REF  -0.08 (-0.15, -0.01) ****** | REF  -0.09 (-0.15, -0.03)******* |
| **Age group (years)**  18-24  25-34  35-44  45-54  55-64  65-74  75+ | 577.6 (122.4-18397.0)  479.6 (104.0-13554.5)  4940.0 (211.5-15636.0)  806.1 (151.8-2959.0)  990.1 (242.8-2568.5)  616.4 (223.4-1963.0)  830.3 (235.9-2029.5) | 990.9 (739.1, 1328.4)  756.1 (608.5, 939.5)  1693.5 (1303.2, 2200.5)  509.2 (335.0, 774.0)  571.9 (473.1, 691.4)  560.7 (508.5, 618.2)  643.8 (488.4, 848.6) | 0.19 (-0.01, 0.38)  0.07 (-0.11, 0.25)  0.42 (0.23, 0.61)*******  -0.10 (-0.33, 0.13)  -0.05 (-0.24, 0.13)  -0.06 (-0.24, 0.12)  REF | 0.45 (0.27, 0.64)*******  0.24 (0.07, 0.41)******  0.36 (0.19, 0.53)*******  0.13 (-0.07, 0.34)  0.03 (-0.13, 0.19)  -0.10 (-0.25, 0.05)  REF |
| **Ethnicity**  White  Mixed  Asian  Black  Other | 805.6 (167.2-4081.0)  583.8 (152.5-13347.0)  1677.0 (227.0-10762.5)  752.7 (190.9-3751.0)  750.8 (79.9-6242.0) | 699.5 (644.7, 758.9)  709.6 (336.8, 1495.0)  1223.4 (818.1, 1829.5)  720.7 (388.5, 1336.8)  462.0 (165.8, 1287.4) | REF   0.01 (-0.27, 0.28)   0.24 (0.07, 0.42)******  0.01 (-0.24, 0.27)  -0.18 (-0.53, 0.17) | REF  -0.09 (-0.32, 0.15)  0.13 (-0.02, 0.28)  0.38 (0.16, 0.60)*******  -0.18 (-0.48, 0.12) |
| **History of COVID-19**  Positive PCR test  Suspected by doctor  Suspected by respondent  No | 6489.0 (749.5-17169.0)  7152.0 (594.4-25000.0)  2314.0 (220.3-15372.0)  535.7 (140.4-2351.5) | 2941.4 (2436.6, 3550.7)  2642.1 (1286.4, 5426.8)  1223.1 (941.6, 1588.7)  493.0 (452.5, 537.2) | 0.78 (0.68, 0.87)*******  0.73 (0.45, 1.0)*******  0.39 (0.30, 0.49)*******  REF | 1.2 (1.1, 1.3)*******  1.0 (0.77, 1.3)*******  0.63 (0.53, 0.72)******  REF |
| **No. of pre-existing health conditions**  >1  1  0 | 523.9 (114.9-2348.0)  714.9 (175.0-3095.0)  1010.0 (194.3-6467.0) | 433.3 (359.8, 521.9)  645.0 (555.6, 748.9)  879.8 (792.6, 976.6) | -0.31 (-0.40, -0.22) *******  -0.13 (-0.22, -0.05) ******  REF | -0.30 (-0.38, -0.23)*******  -0.10 (-0.17, -0.03) ******  REF |
| **Vaccine status**  0  1  2 | 241.9 (20.5-989.7)  333.0 (104.0-15407.0)  1102.0 (287.4-3551.0) | 98.1 (72.4, 133.0)  938.9 (787.1, 1120.1)  953.8 (882.4, 1031.0) | REF  0.98 (0.87, 1.1)*******  0.99 (0.89, 1.1)******* | REF 1.1 (1.0, 1.2)*******  1.7 (1.6, 1,8)******* |
| **Vaccine type (N=2430)**  Pfizer-BioNTech  AstraZeneca  Moderna | 2661.0 (295.0-9918.0)  376.8 (142.1-872.4)  4642.0 (273.9-23049.0) | 1697.0 (1540.4, 1869.6)  344.4 (313.9, 377.8)  2127.8 (1362.8, 3322.2) | REF  -0.69 (-0.76, -0.63)*******  0.10 (-0.07, 0.27) | REF  -0.73 (-0.79, -0.67)*******  0.11 (-0.18, 0.40) |
| **Time since second vaccination (N=2430) (weeks)**  0-3  4-12  13-23  24+ | 12626.0 (4812.0-23190.0)  935.4 (334.0-2848.5)  869.1 (246.9-2480.0)  372 (229.1-655.3) | 6443.4 (5303.4, 7828.5)  973.3 (804.6, 1177.3)  682.2 (625.9, 743.6)  413.0 (299.1, 570.2) | REF  -0.82 (-0.95, -0.69)*******  -0.98 (-1.1, -0.88)*******  -1.2 (-1.5, -0.92)******* | REF  -0.07 (-0.22, 0.08)  -0.27 (-0.41, -0.13)*******  -0.96 (-1.2, -0.70)******* |

Unadjusted regression co-efficient obtained from univariable linear regression for the covariate of interest. Adjusted regression co-efficient obtained from multivariable linear regression adjusting for age, sex, ethnicity, no. of pre-existing health conditions, history of COVID-19 infection and COVID-19 vaccination status. *p<·05, **p<·01, ***p<·001

After adjustment for other demographic and clinical characteristics, higher antibody titres were observed in females compared with males (GMT 768.9 U ml^−1^ (95% CI 694.9, 850.6) vs. 643.8 U ml^−1^ (95% CI 568.6, 728.9)) (Supplementary Table S3). Participants aged 18-44 years had higher antibody titres compared with older participants. Higher antibody titres were also observed in those of Black ethnicity compared to white, and those reporting no pre-existing health conditions (Supplementary Table S3). Previously infected participants and vaccinated participants had higher antibody titres compared with previously uninfected participants and unvaccinated participants, respectively (Supplementary Table S3). Higher antibody titres were observed in those vaccinated with two doses of BNT162b2 mRNA (Pfizer/BioNTech) or mRNA-1273 (Moderna) compared with those vaccinated with two doses of ChAdOx1 nCov-19 (Oxford/AstraZeneca) (GMT 1697.0 U ml^−1^ (95% CI 1540.4, 1869.6) vs. 2127.8 U ml^−1^ (95% CI 1362.8, 3322.2) vs. 344.4 U ml^−1^ (95% CI 313.9, 377.8), respectively. Participants who were within 12 weeks of their second vaccine dose had higher antibody titres compared with participants with a longer time interval since completing their 2-dose primary course (Supplementary Table S3).

Our study provides further evidence for the association between demographic characteristics, history of COVID-19 and vaccination status with levels of SARS-CoV-2 antibody, in keeping with previous studies (1-5). The findings of lower antibody titres after ChAdOx1 nCov-19 vaccination, when compared with BNT162b2 mRNA, and declining titres with increasing time since vaccination are consistent with other real-world data (1, 5). Our findings are also consistent with patterns in SARS-CoV-2 antibody prevalence seen in the national REACT-2 study using the Fortress LFIA, where the highest prevalence was found in females, people of non-white ethnicity, younger adults and those with previous infection (6-8).

**REFERENCES**

1.	Steensels D, Pierlet N, Penders J, Mesotten D, Heylen L. Comparison of SARS-CoV-2 Antibody Response Following Vaccination With BNT162b2 and mRNA-1273. JAMA. 2021;326(15):1533-5.

2.	Zeng F, Dai C, Cai P, Wang J, Xu L, Li J, et al. A comparison study of SARS-CoV-2 IgG antibody between male and female COVID-19 patients: A possible reason underlying different outcome between sex. J Med Virol. 2020;92(10):2050-4.

3.	Wei J, Matthews PC, Stoesser N, Maddox T, Lorenzi L, Studley R, et al. Anti-spike antibody response to natural SARS-CoV-2 infection in the general population. Nature Communications. 2021;12(1):6250.

4.	Pellini R, Venuti A, Pimpinelli F, Abril E, Blandino G, Campo F, et al. Initial observations on age, gender, BMI and hypertension in antibody responses to SARS-CoV-2 BNT162b2 vaccine. EClinicalMedicine. 2021;36:100928-.

5.	Wei J, Pouwels KB, Stoesser N, Matthews PC, Diamond I, Studley R, et al. Antibody responses and correlates of protection in the general population after two doses of the ChAdOx1 or BNT162b2 vaccines. Nature Medicine. 2022.

6.	Ward H, Cooke GS, Atchison C, Whitaker M, Elliott J, Moshe M, et al. Prevalence of antibody positivity to SARS-CoV-2 following the first peak of infection in England: Serial cross-sectional studies of 365,000 adults. Lancet Reg Health Eur. 2021;4:100098.

7.	Ward H, Atchison C, Whitaker M, Ainslie KEC, Elliott J, Okell L, et al. SARS-CoV-2 antibody prevalence in England following the first peak of the pandemic. Nature Communications. 2021;12(1):905.

8.	Ward H, Whitaker M, Flower B, Tang SN, Atchison C, Darzi A, et al. Population antibody responses following COVID-19 vaccination in 212,102 individuals. Nature Communications. 2022;13(1):907.
