## Supplemental Figure 1 for "Validity of self-testing at home with rapid SARS-CoV-2 antibody detection by lateral flow immunoassay"

**Supplementary figure S1.** The conversion of SARS-CoV-2 live virus neutralisation titres to BAU/ mL by titration of a WHO antibody reference standard (20/150; anti-Spike IgG 832 BAU/ mL) (1)

| **SARS-CoV-2 NT_50_** | **BAU/ mL** |
| --- | --- |
| 0 | 0 |
| 7.1 | 18 |
| 10 | 26 |
| 14.1 | 37 |
| 20 | 52 |
| 28.3 | 74 |
| 40 | 104 |
| 56.6 | 147 |
| 80 | 208 |
| 113.1 | 294 |
| 160 | 416 |
| 226.3 | 588 |
| 320 | 832 |
| 452.5 | 1177 |
| 640 | 1664 |
| 905.1 | 2353 |
| 1280 | 3328 |
| 1810.2 | 4707 |
| 2560 | 6656 |
| 3620.4 | 9413 |
| 5120 | 13312 |
| 7240.8 | 18826 |
| 10240 | 26624 |
| 14481.5 | 37652 |
| 20480 | 53248 |

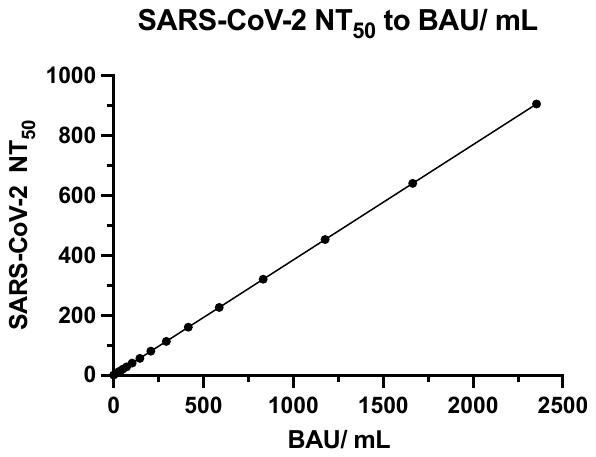

The conversion of SARS-CoV-2 live neutralisation titres to BAU/mL by titration of a WHO antibody reference standard (20/150; anti-Spike IgG 832 BAU/ mL) (1)

**References**

1.	Mattiuzzo G, Bentley EM, Hassall M, Routley S, Richardson S, Bernasconi V, et al. WHO/BS.2020.2403 Establishment of the WHO International Standard and Reference Panel for anti-SARS-CoV-2 antibody2020; (20 April 2022). Available from: <https://www.nibsc.org/documents/ifu/20-268.pdf>.
